# Surgical interventions in idiopathic intracranial hypertension – a comprehensive multi-center study of outcome and the role of treatment indication

**DOI:** 10.1101/2025.07.14.25331492

**Authors:** Gabriel Bsteh, Nadja Skadkær Hansen, Sina Zaic, Steffen Hamann, Johanne Juhl Korsbæk, Nik Krajnc, Stefan Macher, Laleh Dehghani Molander, Klaus Novak, Therese Wallentin Steenfoss, Marianne Wegener, Berthold Pemp, Rigmor Hoejland Jensen, Dagmar Beier

## Abstract

**Background:** Surgical intervention is recommended in idiopathic intracranial hypertension (IIH) for fulminant or treatment-refractory cases, yet data on outcomes remain limited, particularly regarding indication-specific effects. This study evaluated outcomes and indications for surgery in IIH, aiming to identify predictors of favorable or adverse results.

**Methods:** A retrospective multi-center study was conducted by the Danish-Austrian IIH Consortium (DASH-IIH). Databases from three centers (Vienna, Odense, Copenhagen) were screened for people with IIH (pwIIH) fulfilling revised Friedman criteria who underwent surgery and had ≥6 months of follow-up. Outcomes at six months included visual function, headache improvement (≥50%), papilledema resolution, and severe adverse events. Multivariable regression was used to adjust for confounders.

**Results:** Of 1310 pwIIH, only 3.6% required surgery overall. Thirty-six pwIIH were included (100% female; mean age 32.5 years; median BMI 37.0; median CSF opening pressure 41 cmH□O). Of these, 27 (75%) underwent CSF diversion and 9 (25%) optic nerve sheath fenestration (ONSF). The primary indication for surgery was acute visual deterioration in 83.3% and refractory headache in 16.7%.

Visual function improved in 41.7%, papilledema resolved in 89.7%, and headache improved in 30.6%. No significant differences in outcomes were found between CSF diversion and ONSF. Importantly, no visual improvement occurred in cases operated for headache alone, and the odds of headache improvement were significantly lower in this group (OR 0.11, p=0.012).

**Conclusion:** CSF diversion and ONSF are effective in IIH with acute visual threat, improving vision and, to a much lesser extent, headache. Refractory headache alone appears insufficient indication for surgical intervention.

**What is already known on this topic:** Surgical interventions such as cerebrospinal fluid (CSF) diversion and optic nerve sheath fenestration (ONSF) are established treatments for fulminant or treatment-refractory idiopathic intracranial hypertension (IIH), particularly when vision is acutely threatened. However, previous studies are limited to small, single-center case series and lack robust analyses of how treatment indication – particularly refractory headache versus visual deterioration – affects clinical outcomes.

**What this study adds:** This multi-center, population-based study document that only a selected subset of IIH patients need surgery and provides the first comparative analysis of surgical outcomes in IIH stratified by treatment indication. It shows that both CSF diversion and ONSF are similarly effective in preserving vision when performed for acute visual deterioration. In contrast, surgical intervention for refractory headache alone improves headache markedly less and has no visual benefit, suggesting that headache alone may not justify invasive treatment.

**How this study might affect research, practice, or policy:** These findings reinforce the importance of precise, indication-based patient selection for surgical treatment in IIH. They support current consensus guidelines recommending multidisciplinary evaluation and suggest that refractory headache, in the absence of visual threat, should not be considered a standalone surgical indication. The results may inform clinical decision-making and prioritize non-surgical management for headache-dominant IIH. Future research should investigate tailored treatment strategies and alternative therapies for this subgroup.

## Introduction

Idiopathic intracranial hypertension (IIH) – formerly known as pseudotumor cerebri or benign intracranial hypertension – is a syndrome characterized by increased intracranial pressure (ICP) of unknown etiology [1]. Although IIH is considered rare in the general population, it predominantly affects obese women of childbearing age, and its incidence is rising amid the global obesity pandemic [2, 3]. Beyond the primary risk of visual impairment, long-term outcomes in people with IIH (pwIIH) are often driven by disabling, chronic headaches [4]. Once the diagnosis is established, treatment goals focus on preserving vision, alleviating headaches, and improving quality of life while carefully managing treatment side effects and risks [5]. Given the complexity of diagnosing, treating, and monitoring IIH, international consensus guidelines recommend that care should be provided in specialized, multidisciplinary centers offering comprehensive monitoring [6–11]. Although most pwIIH can be managed conservatively, invasive surgical intervention should be immediately considered in case of acute and severe visual deterioration [5, 12, 13].

Invasive treatment options include cerebrospinal fluid (CSF) diversion procedures – such as ventriculoperitoneal / -atrial (VP / VA) shunting – and optic nerve sheath fenestration (ONSF). Meta-analyses indicate that both provide improvement of vision in approximately 50% of pwIIH, though CSF diversion procedures seem to be associated with higher revision rates for complications (typically device associated), whereas ONSF seems to have a higher rate of failure due to re-occlusion of the nerve sheath [14–17].

However, current evidence exclusively stems from case reports or single-center case series characterized by small sample sizes and highly heterogeneous outcome definitions – especially regarding headache outcomes [14, 15, 17, 18]. Moreover, many studies lack a comprehensive evaluation of the pre- and post-surgical course, often omitting key clinical parameters. Notably, the clinical indication for surgical interventions, which could significantly influence outcomes, have not been adequately addressed in previous analyses.

In this comprehensive, multi-center study, we aim to bridge these gaps by systematically evaluating both the outcomes of surgical interventions in IIH and the treatment indication that precede these procedures.

## Methods

### Patient population

This study was designed as a retrospective multi-center cohort study by the Danish Austrian IIH Consortium (DASH-IIH) comprising the specialized multidisciplinary IIH centers at the Medical University of Vienna (Austria), the Copenhagen University Hospital – Rigshospitalet (Denmark) and the Odense University Hospital (Denmark). We included all patients with definite IIH according to the modified Friedman criteria aged ≥18 years who i) underwent a surgical intervention for IIH (VP shunting, VA shunting, ONSF) between 01-JAN-2014 and 01-JUN-2024 (in Copenhagen 01-JAN-2016 to 01-JUN-2024 due to unavailability of data beforehand), and ii) had clinical and paraclinical data available within 30 days before surgical intervention (at least one complete clinical and paraclinical assessment) and at least 6 months after the intervention [19].

### Ophthalmological assessment

Best corrected visual acuity was determined by Snellen tables at distance after subjective refraction by minimum angle of resolution (logMAR). Static threshold perimetry was performed using 30-2 (Vienna: Humphrey, Copenhagen: Octopus) or 24-2 (Odense: Humphrey, Carl Zeiss) standard protocols, quantifying the mean deviation in decibels (dB) in perimetric mean deviation (MD) of all test locations compared to age-matched controls (9,12). All perimetric results were interpreted by the assessing neuroophthalmologist differentiating real visual field defects from artefacts caused by insufficient cooperation to the investigation. Visual impairment was defined as a visual acuity deviation of ≥0.1 logMAR and/or <-2.0 dB in MD [20]. Optic disc evaluation was performed using standardized ophthalmoscopy and/or fundus photography, and the Frisén grading scale was used to rate papilledema severity, categorizing the swelling of optic discs from grade 0 (no papilledema) to grade 5 (severe papilledema) (12). If the eyes were differentially affected, the grading of the most severely affected eye was used. The assessing neuroophthalmologist differentiated papilledema from pseudopapilledema and swelling of the optic disc due to other causes than elevated ICP.

OCT imaging was done without pupil dilation using spectral-domain OCT (Spectralis OCT, Heidelberg Engineering, Heidelberg, Germany), adhering to the OSCAR-IB quality control criteria and describing findings in accordance with the APOSTEL criteria (13,14). For peripapillary retinal nerve fiber layer (pRNFL) thickness measurement, a 12° ring scan centered on the optic nerve head was used (15). Macular ganglion cell layer (GCL) volume was measured in both eyes of each pwIIH using a macular volume scan corresponding to the 6 mm ring area of the circular grid defined by the Early Treatment Diabetic Retinopathy Study (16). Image processing was semiautomated using the built-in proprietary software for automated layer segmentation and manual correction of obvious errors. Measurements of worse eyes were used for statistical analysis, i.e., higher pRNFL thickness as a marker of papilledema and lower GCL volume as a marker of neuroaxonal loss.

### Headache assessment

Headache outcome was retrieved from documented headache diaries obtaining headache frequency (determined by monthly headache days [MHD]) and headache severity (on the numerical rating scale [NRS]). Adverse events were retrieved from medical records.

### Endpoints

The primary endpoint was set as visual improvement 6 months (+/-30 days) after surgical intervention (M6) defined as improvement of best corrected visual acuity by ≥0.2 logMAR and/or by ≥2.0 dB in perimetric MD compared to baseline.

Secondary endpoints included headache improvement (defined as ≥50% reduction in headache severity and/or headache frequency compared to baseline) at M6, resolution of papilledema (defined as Frisén grade ≤1 at M6), loss of GCL volume (defined as a loss of ≥0.02 mm^3^ at M6 compared to baseline), visual worsening at M6 (defined as worsening of best corrected visual acuity by ≥0.2 logMAR and/or by ≥2.0 dB in perimetric MD compared to baseline), and occurrence of severe adverse events (SAE; defined as Common Terminology Criteria for Adverse Events (CTCAE, v6.0) grade ≥3) [21].

### Predictors of main interest

The first main predictor of interest was the type of surgical intervention classified as either CSF diversion procedures (comprising all types of CSF shunting) or ONSF.

The second main predictor of interest was the indication for surgical intervention, classified as either acute threat of visual deterioration or refractory headache. If both were present, a classification as acute threat of visual deterioration was made.

### Data curation and data analysis

The data relevant to this study were extracted from local databases at three centers in Vienna (VIIH database), Odense and Copenhagen, which contain data collected both retrospectively and prospectively with thorough quality control established [8–10].

### Statistical analyses

Statistical analysis was performed using R-Statistical Software (Version 4.4.2). After description of primary endpoints according to different surgical interventions, systematic univariable analyses of baseline parameters for association with endpoints were performed using the chi-square test, Fisher exact test, Mann-Whitney U test or independent t-test (with Welch correction for unequal standard deviations between the groups) as appropriate. Univariable correlation analyses were calculated using Pearson or Spearman-rho tests, depending on the presence of a normal distribution.

To investigate predictors of clinical outcome, multivariable analyses were performed using binary-logistic bias-reduced logistic regression models according to Firth (a method based on penalized likelihood, which increases the efficiency of the estimators in logistic regression models with small samples; R package “logistf”, version 1.24.1) with primary and secondary endpoints as the dependent variable and predictors of main interest as the independent variables (CSF diversion vs. ONSF; acute threat of visual deterioration vs. refractory headache acute visual) [22]. Corrected Akaike information criterion (AICc) was used to select the best-fitting model for adjustment from a predefined set of known relevant covariables (for visual improvement/worsening: sex, age, time since diagnosis, body mass index (BMI), visual impairment at baseline, CSF opening pressure (CSF-OP), Frisén scale at baseline [Frisén_Base_], GCL volume at baseline [GCL_Base_], pRNFL at baseline [pRNFL_Base_]; for headache improvement: sex, age, time since diagnosis, BMI, CSF-OP, MHD/headache severity at baseline; for resolution of papilledema: sex, age, time since diagnosis, BMI, CSF-OP, Frisén_Base_, pRNFL_Base_; for loss in GCL volume: sex, age, BMI, CSF-OP, Frisén_Base_, GCL_Base_, pRNFL_Base_; for SAE: sex, age, time since diagnosis) and all other variables displaying an association with the respective outcome parameter p<0.2 in univariable analyses [23]. The robustness of all regression models to unidentified confounding factors (bias) was quantified using the Rosenbaum sensitivity test according to Hodges-Lehmann Gamma [24]. We tested all variables for normal distribution by Lilliefors-test and for collinearity by variance inflation factor (VIF) and excluded all variables from the regression analyses if the VIF was >2.0 corresponding to an *R^2^* of ≥0.50. Missing values were treated by multiple (20-fold) imputation using the MNAR (Missing not at Random) approach with pooling of estimates according to Rubin’s rules [25]. Significance level was set at a two-sided p-value <0.05.

### Standard Protocol Approvals, Registrations, and Patient Consents

The study was approved by the ethics committee of the Medical Universities of Vienna (ethics approval number: 2216/2020), the Region of Southern Denmark (journal number: 24/17408, approval covers all Danish sites) and the Danish Data Protection Agency (journal number: 24/21145). As this was a retrospective study, the need for written informed consent from study participants was waived by the ethics committee and the Region of Southern Denmark. This study adheres to the reporting guidelines outlined within the ‘Strengthening the Reporting of Observational Studies in Epidemiology (STROBE) Statement.

### Data Availability Statement

Individual, de-identified participant data can be shared with qualified researchers who provide a methodologically sound proposal and approval by the responsible ethics committees. It is a legal requirement that a data processing agreement is signed and approved by the data protection office in the Region of Southern Denmark as well as the data-clearing committee of the Medical Universities Vienna. Raw imaging data are shared locally for technical and legal reasons. Data are available two years after publication. Proposals should be directed to the corresponding author.

## Results

The final study cohort comprised 36 pwIIH (Vienna: 14, Odense: 12, Copenhagen: 10). The detailed inclusion/exclusion process is depicted in Figure 1. Overall, the proportion of pwIIH requiring surgical intervention within the setting of these specialized multidisciplinary IIH centers was 3.6%.

**Figure 1.**
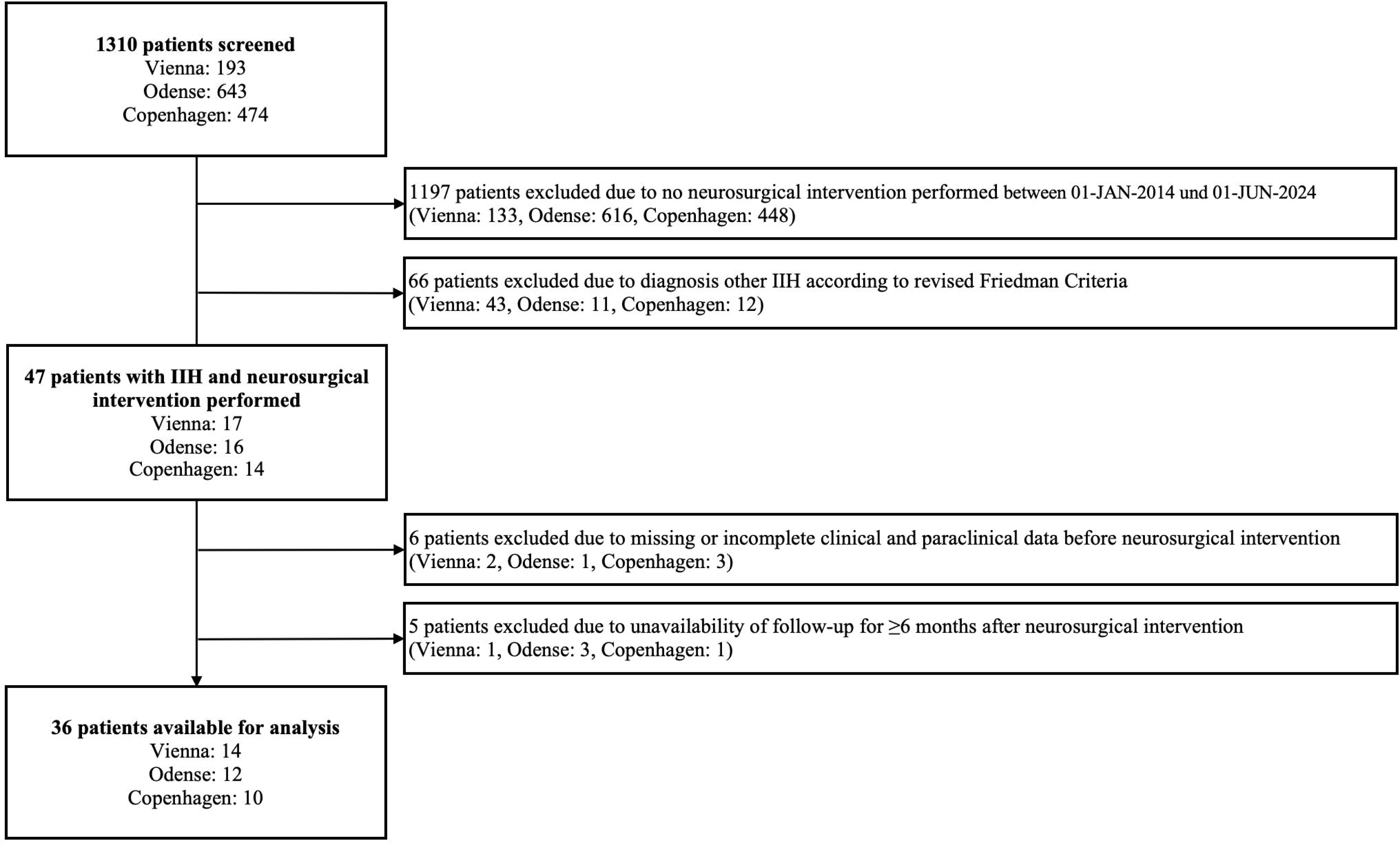
Flow chart of inclusion/exclusion process.

Baseline characteristics of the study cohort before surgical intervention are shown in Table 1. Overall, 27 pwIIH received CSF diversion procedures, with 25 (69.4%) VP and 2 (5.6%) VA shunting, whereas 9 (25.0%) received ONSF (Figure 2a).

**Figure 2.**
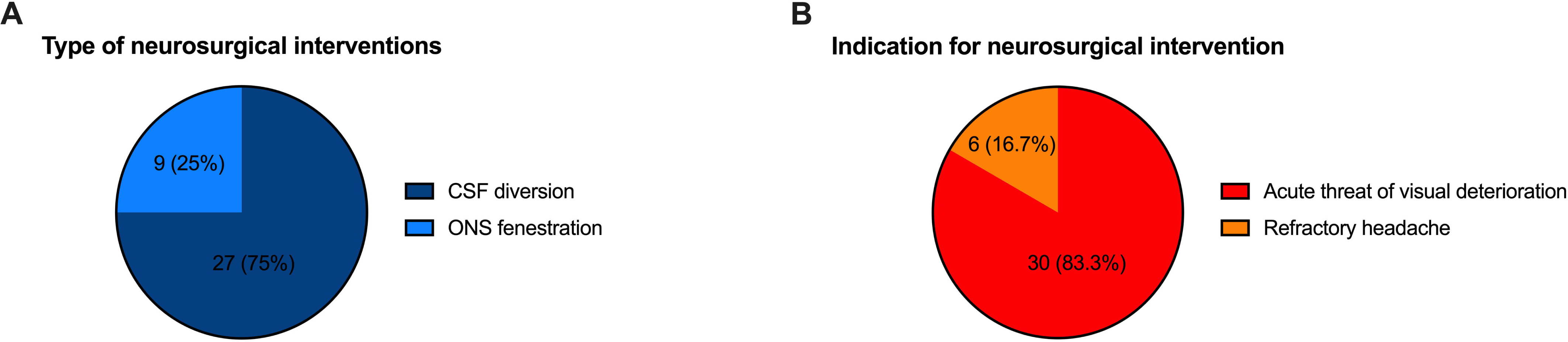
Overall distribution of type and indication for surgical interventions.

**Table 1.** Cohort characteristics before surgical intervention.

|  |  |
| --- | --- |
|  | (n=36) |
| Females<sup>1</sup> | 36 (100) |
| Age at diagnosis<sup>2</sup> (years) | 32.5 (8.2) |
| Time since diagnosis at surgery<sup>3</sup> (months) | 1.6 (0.2 – 16.4) |
| BMI<sup>3</sup> | 37.0 (32.5 – 45.1) |
| CSF-OP<sup>3</sup> (cm H2O)<sup>3</sup> (cm H2O) | 41.5 (33.5 – 50) |
| Visual impairment<sup>1</sup> | 31 (86.1) |
| Best corrected visual acuity<sup>3</sup> (logMAR, worse eye) | 0.125 (0 – 0.40) |
| Perimetric mean defect<sup>2</sup> (dB, worse eye) | -13.0 (-25.5 – -3.0) |
| Frisén grade<sup>3</sup> | 3 (2 – 4) |
| pRNFL thickness<sup>2</sup> (µm, worse eye) | 228 (133) |
| GCL volume<sup>2</sup> (mm<sup>3</sup>, worse eye)* | 0.99 (0.18) |
| Headache frequency<sup>3</sup> (days per month) | 30 (30 – 30) |
| Headache severity<sup>3</sup> (NRS) | 8 (7 – 10) |
| Acetazolamide dosage<sup>3</sup> (mg/day) | 1500 (1000 – 2250) |
| Number of lumbar punctures before surgery<sup>3</sup> | 2 (1 – 4) |

**Abbreviations:** BMI: Body mass index. dB: decibels. CSF-OP: cerebrospinal fluid opening pressure GCL: ganglion cell layer. logMAR: logarithm of the minimum angle of resolution. pRNFL: peripapillary retinal nerve fiber layer. NRS: numerical rating scale.
<sup>1</sup>absolute number and percentage <sup>2</sup>mean and standard deviation <sup>3</sup>median and inter-quartile range

The primary indication for intervention was an acute threat of visual deterioration in 30 pwIIH (83.3%) and refractory headache in 6 pwIIH (16.7%) (Figure 2b).

The primary endpoint, visual improvement at M6, was observed in 15 pwIIH (41.7%; Figure 3a), while resolution of papilledema occurred in 32 (88.9%; Figure 3b). Headache improvement was observed in 10 pwIIH (30.6%) (Figure 3c). Loss of GCL volume was found in 16.7% (6/16) pwIIH with sufficient data, whereby the mean loss of GCL volume was 0.13mm^3^ (SD 0.17). SAEs were reported in 14 pwIIH (38.9%), with the majority attributed to revisions (12/36, 33.3%) and five patients (13.9%) experienced visual worsening at M6 (Supplemental Table 1).

**Figure 3.**
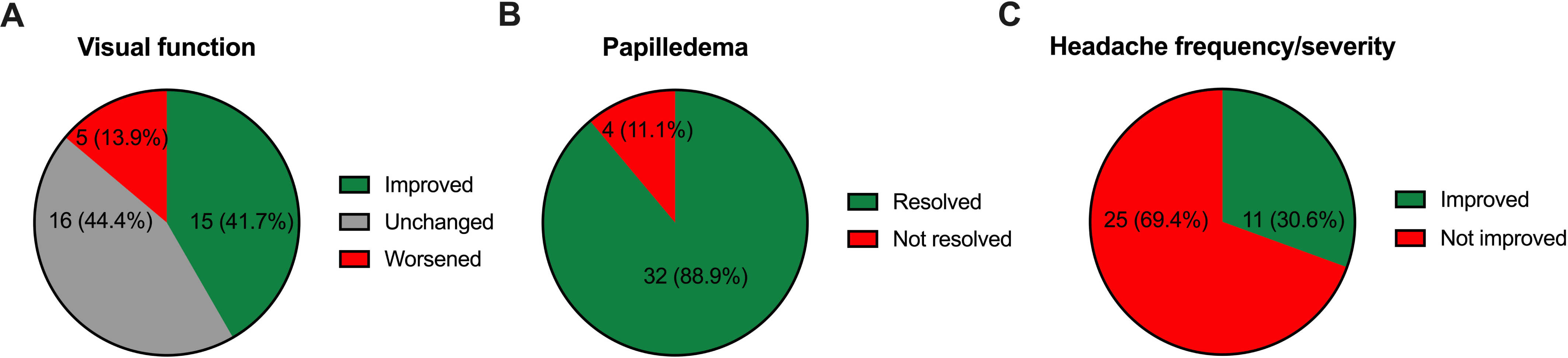
Overall frequency of main outcome measures.

### CSF diversion vs. optic nerve sheath fenestration

In univariable comparisons, we found no significant differences between CSF diversion procedures and ONSF regarding visual improvement (33.3% vs. 66.6%, p=0.122), visual worsening (11.1% vs. 22.2%, p=0.581), papilledema resolution (86.4% vs. 87.5%, p=0.999), headache improvement (35.3% vs. 55.6%, p=0.419), or GCL volume loss (38.5% vs. 33.3%, p=0.999). There were also no significant differences between the groups in any of the baseline characteristics (Supplemental Table 2).

Overall rates of SAEs were comparable between CSF diversion and ONSF (38.5% vs. 44.4%, p=0.526), as well as overall rate of revision surgery (34.6% vs. 33.3%, p=0.639). In ONSF, revisions occurred due to failure to resolve papilledema (33.3% vs. 0% in CSF diversion, p=0.013), while revisions in pwIIH with CSF diversion were exclusively due to dysfunction or shunt infection after initial resolution of papilledema (34.6% vs. 0%, p=0.074).

Multivariable models, adjusted for relevant covariables, confirmed that the type of surgical intervention had no significant impact on any outcome measures (Table 2). While the overall goodness-of-fit of the multivariable models was moderate (Pseudo-R-squared 0.311-0.469), Hodges-Lehmann Gamma indicated robustness to bias by unidentified confounders.

**Table 2.**
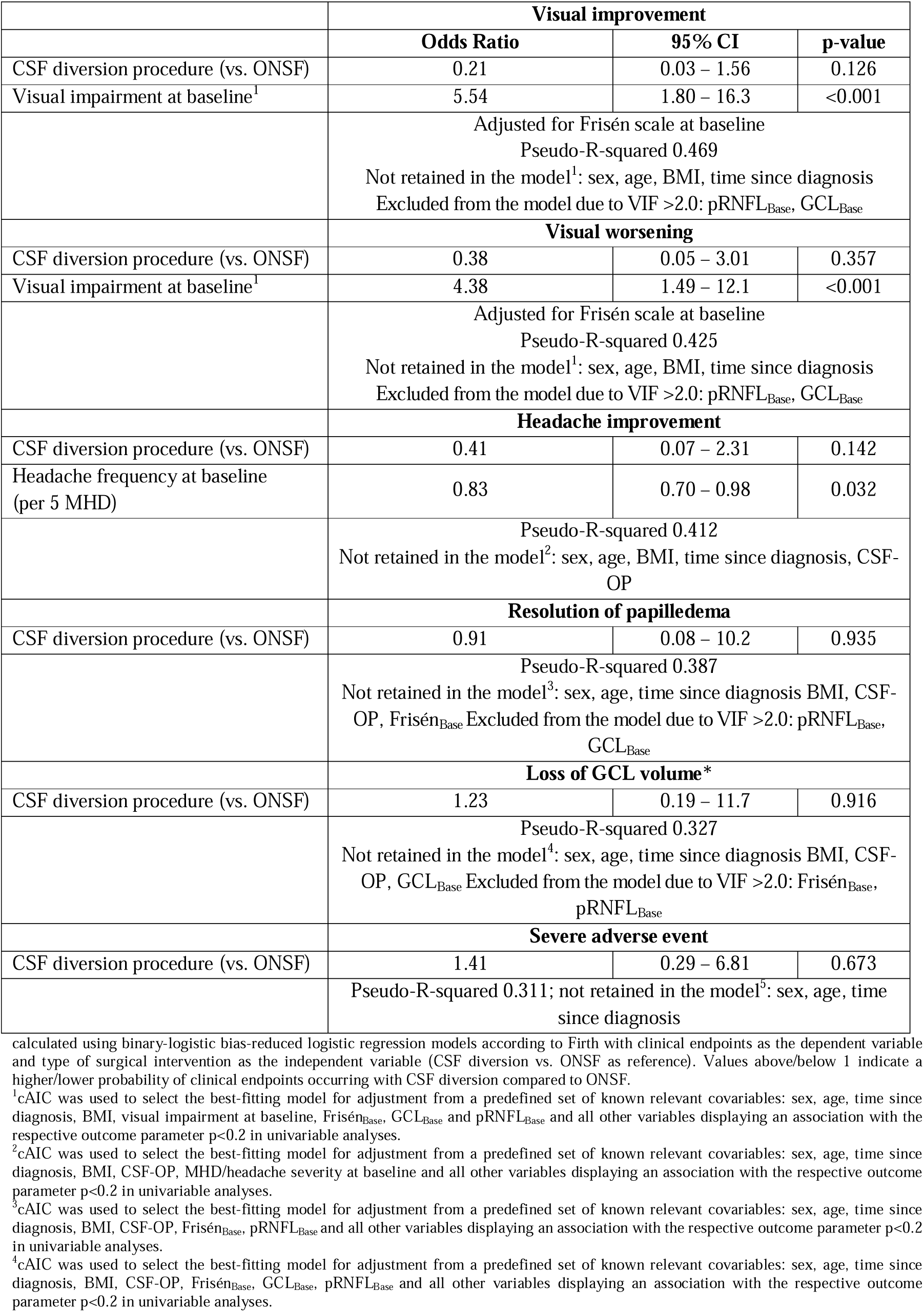
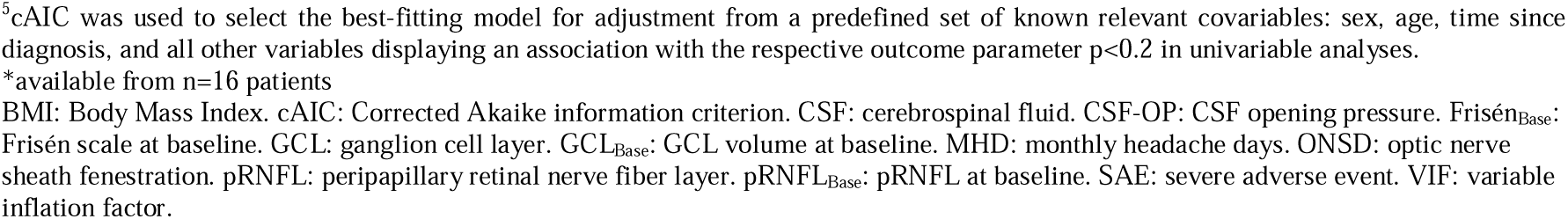
Impact of type of surgical intervention on six-month clinical outcomes in IIH: multivariable analysis.

|  | **Visual improvement** |  |  |
| --- | --- | --- | --- |
|  | **Odds Ratio** | **95% CI** | **p-value** |
| CSF diversion procedure (vs. ONSF) | 0.21 | 0.03 – 1.56 | 0.126 |
| Visual impairment at baseline<sup>1</sup> | 5.54 | 1.80 – 16.3 | <0.001 |
|  | Adjusted for Frisén scale at baseline<br>Pseudo-R-squared 0.469<br>Not retained in the model<sup>1</sup>: sex, age, BMI, time since diagnosis<br>Excluded from the model due to VIF >2.0: pRNFL<sub>Base</sub>, GCL<sub>Base</sub> |  |  |
|  | **Visual worsening** |  |  |
| CSF diversion procedure (vs. ONSF) | 0.38 | 0.05 – 3.01 | 0.357 |
| Visual impairment at baseline<sup>1</sup> | 4.38 | 1.49 – 12.1 | <0.001 |
|  | Adjusted for Frisén scale at baseline<br>Pseudo-R-squared 0.425<br>Not retained in the model<sup>1</sup>: sex, age, BMI, time since diagnosis<br>Excluded from the model due to VIF >2.0: pRNFL<sub>Base</sub>, GCL<sub>Base</sub> |  |  |
|  | **Headache improvement** |  |  |
| CSF diversion procedure (vs. ONSF) | 0.41 | 0.07 – 2.31 | 0.142 |
| Headache frequency at baseline (per 5 MHD) | 0.83 | 0.70 – 0.98 | 0.032 |
|  | Pseudo-R-squared 0.412<br>Not retained in the model<sup>2</sup>: sex, age, BMI, time since diagnosis, CSF-OP |  |  |
|  | **Resolution of papilledema** |  |  |
| CSF diversion procedure (vs. ONSF) | 0.91 | 0.08 – 10.2 | 0.935 |
|  | Pseudo-R-squared 0.387<br>Not retained in the model<sup>3</sup>: sex, age, time since diagnosis BMI, CSF-OP, Frisén<sub>Base</sub> Excluded from the model due to VIF >2.0: pRNFL<sub>Base</sub>, GCL<sub>Base</sub> |  |  |
|  | **Loss of GCL volume\*** |  |  |
| CSF diversion procedure (vs. ONSF) | 1.23 | 0.19 – 11.7 | 0.916 |
|  | Pseudo-R-squared 0.327<br>Not retained in the model<sup>4</sup>: sex, age, time since diagnosis BMI, CSF-OP, GCL<sub>Base</sub> Excluded from the model due to VIF >2.0: Frisén<sub>Base</sub>, pRNFL<sub>Base</sub> |  |  |
|  | **Severe adverse event** |  |  |
| CSF diversion procedure (vs. ONSF) | 1.41 | 0.29 – 6.81 | 0.673 |
|  | Pseudo-R-squared 0.311; not retained in the model<sup>5</sup>: sex, age, time since diagnosis |  |  |

calculated using binary-logistic bias-reduced logistic regression models according to Firth with clinical endpoints as the dependent variable and type of surgical intervention as the independent variable (CSF diversion vs. ONSF as reference). Values above/below 1 indicate a higher/lower probability of clinical endpoints occurring with CSF diversion compared to ONSF.
<sup>1</sup>cAIC was used to select the best-fitting model for adjustment from a predefined set of known relevant covariables: sex, age, time since diagnosis, BMI, visual impairment at baseline, Frisén<sub>Base</sub>, GCL<sub>Base</sub> and pRNFL<sub>Base</sub> and all other variables displaying an association with the respective outcome parameter p<0.2 in univariable analyses.
<sup>2</sup>cAIC was used to select the best-fitting model for adjustment from a predefined set of known relevant covariables: sex, age, time since diagnosis, BMI, CSF-OP, MHD/headache severity at baseline and all other variables displaying an association with the respective outcome parameter p<0.2 in univariable analyses.
<sup>3</sup>cAIC was used to select the best-fitting model for adjustment from a predefined set of known relevant covariables: sex, age, time since diagnosis, BMI, CSF-OP, Frisén<sub>Base</sub>, pRNFL<sub>Base</sub> and all other variables displaying an association with the respective outcome parameter p<0.2 in univariable analyses.
<sup>4</sup>cAIC was used to select the best-fitting model for adjustment from a predefined set of known relevant covariables: sex, age, time since diagnosis, BMI, CSF-OP, Frisén<sub>Base</sub>, GCL<sub>Base</sub>, pRNFL<sub>Base</sub> and all other variables displaying an association with the respective outcome parameter p<0.2 in univariable analyses.
<sup>5</sup>cAIC was used to select the best-fitting model for adjustment from a predefined set of known relevant covariables: sex, age, time since diagnosis, and all other variables displaying an association with the respective outcome parameter $p < 0.2$ in univariable analyses.
\*available from n=16 patients
BMI: Body Mass Index. cAIC: Corrected Akaike information criterion. CSF: cerebrospinal fluid. CSF-OP: CSF opening pressure. Frisén<sub>Base</sub>: Frisén scale at baseline. GCL: ganglion cell layer. GCL<sub>Base</sub>: GCL volume at baseline. MHD: monthly headache days. ONSD: optic nerve sheath fenestration. pRNFL: peripapillary retinal nerve fiber layer. pRNFL<sub>Base</sub>: pRNFL at baseline. SAE: severe adverse event. VIF: variable inflation factor.

### Indication for surgery as a predictor of outcome

Compared to interventions for an acute threat of visual deterioration, surgical procedures performed for refractory headache achieved a significantly lower rate of visual improvement (0% vs. 50.0%, p=0.030). In contrast, there were no significant differences between the groups regarding visual worsening (0% vs. 16.7%, p=0.564), papilledema resolution (100% vs. 86.7%, p=0.999), headache improvement (16.7% vs. 30.0%, p=0.656), GCL volume loss (0% vs. 46.2%, p=0.250), or SAEs (33.3% vs. 40.0%, p=0.999). Time since diagnosis was significantly longer when surgery was indicated for refractory headache compared to acute threat of visual deterioration (25.1 vs. 0.4 months, p=0.002). PwIIH receiving surgery for acute threat of visual deterioration had significantly more visual impairment (100% vs. 16.7%, p<0.001), higher Frisén grade (median 4 vs. 1, p=0.003), higher CSF-OP (43 vs 28 cmH2O, p=0.011), and thicker pRNFL (262 µm vs. 118µm, p<0.001) (Table 3).

**Table 3.**
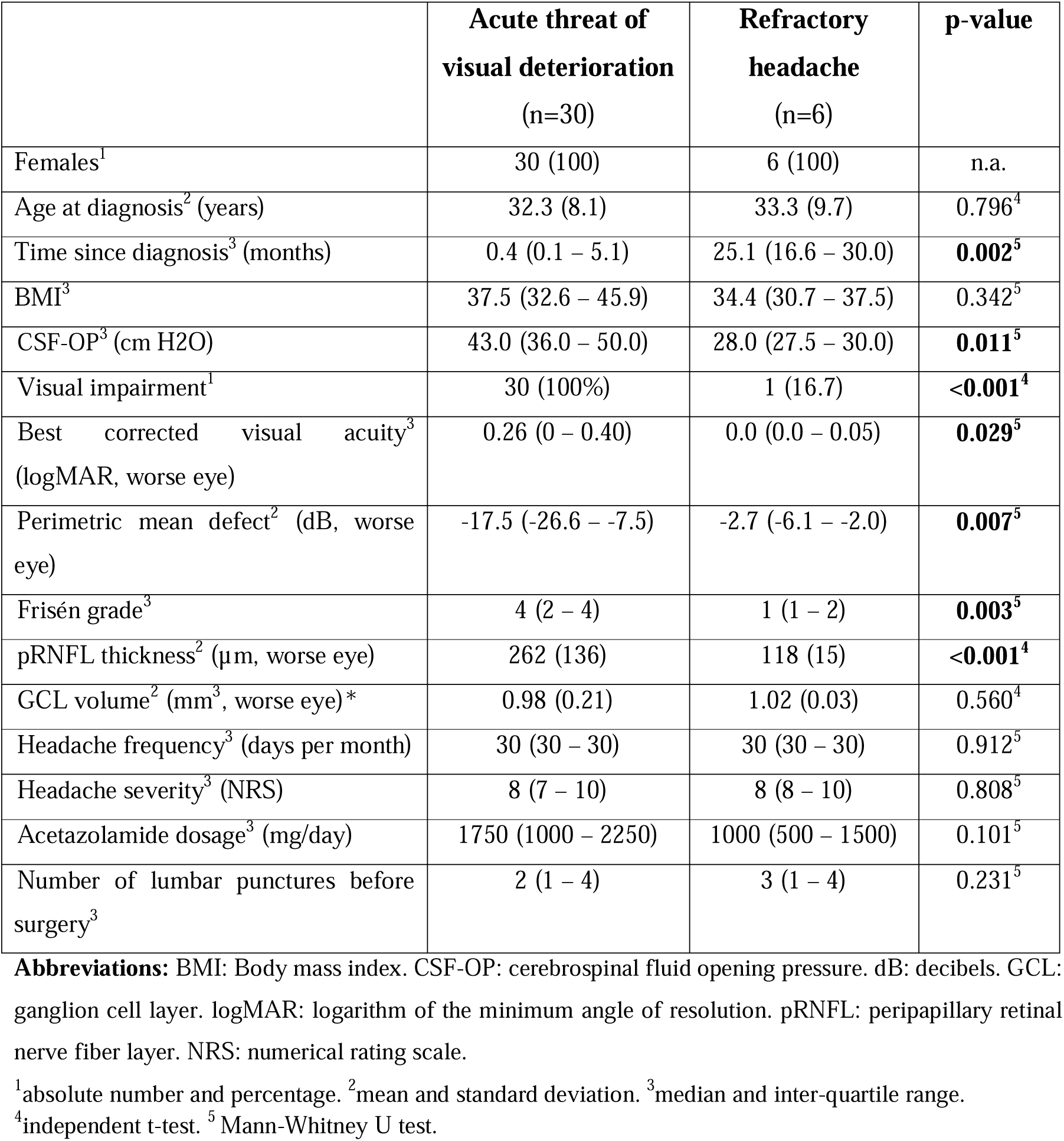
Baseline characteristics stratified by indication for surgical intervention.

|  | **Acute threat of**<br>**visual deterioration**<br>(n=30) | **Refractory**<br>**headache**<br>(n=6) | **p-value** |
| --- | --- | --- | --- |
| Females<sup>1</sup> | 30 (100) | 6 (100) | n.a. |
| Age at diagnosis<sup>2</sup> (years) | 32.3 (8.1) | 33.3 (9.7) | 0.796<sup>4</sup> |
| Time since diagnosis<sup>3</sup> (months) | 0.4 (0.1 – 5.1) | 25.1 (16.6 – 30.0) | **0.002<sup>5</sup>** |
| BMI<sup>3</sup> | 37.5 (32.6 – 45.9) | 34.4 (30.7 – 37.5) | 0.342<sup>5</sup> |
| CSF-OP<sup>3</sup> (cm H2O) | 43.0 (36.0 – 50.0) | 28.0 (27.5 – 30.0) | **0.011<sup>5</sup>** |
| Visual impairment<sup>1</sup> | 30 (100%) | 1 (16.7) | **<0.001<sup>4</sup>** |
| Best corrected visual acuity<sup>3</sup><br>(logMAR, worse eye) | 0.26 (0 – 0.40) | 0.0 (0.0 – 0.05) | **0.029<sup>5</sup>** |
| Perimetric mean defect<sup>2</sup> (dB, worse<br>eye) | -17.5 (-26.6 – -7.5) | -2.7 (-6.1 – -2.0) | **0.007<sup>5</sup>** |
| Frisén grade<sup>3</sup> | 4 (2 – 4) | 1 (1 – 2) | **0.003<sup>5</sup>** |
| pRNFL thickness<sup>2</sup> (µm, worse eye) | 262 (136) | 118 (15) | **<0.001<sup>4</sup>** |
| GCL volume<sup>2</sup> (mm<sup>3</sup>, worse eye)* | 0.98 (0.21) | 1.02 (0.03) | 0.560<sup>4</sup> |
| Headache frequency<sup>3</sup> (days per month) | 30 (30 – 30) | 30 (30 – 30) | 0.912<sup>5</sup> |
| Headache severity<sup>3</sup> (NRS) | 8 (7 – 10) | 8 (8 – 10) | 0.808<sup>5</sup> |
| Acetazolamide dosage<sup>3</sup> (mg/day) | 1750 (1000 – 2250) | 1000 (500 – 1500) | 0.101<sup>5</sup> |
| Number of lumbar punctures before<br>surgery<sup>3</sup> | 2 (1 – 4) | 3 (1 – 4) | 0.231<sup>5</sup> |

**Abbreviations:** BMI: Body mass index. CSF-OP: cerebrospinal fluid opening pressure. dB: decibels. GCL: ganglion cell layer. logMAR: logarithm of the minimum angle of resolution. pRNFL: peripapillary retinal nerve fiber layer. NRS: numerical rating scale.
<sup>1</sup>absolute number and percentage. <sup>2</sup>mean and standard deviation. <sup>3</sup>median and inter-quartile range.
<sup>4</sup>independent t-test. <sup>5</sup> Mann-Whitney U test.

**Table 4.** Impact of indication on six-month clinical outcomes after surgical intervention in IIH: multivariable analysis.

|  | **Visual improvement** |  |  |
| --- | --- | --- | --- |
|  | **Odds Ratio** | **95% CI** | **p-value** |
| Indication: refractory headache<br>(vs. acute threat of visual deterioration) | Not calculable because no cases of visual improvement were observed in the refractory headache group. |  |  |
|  | **Visual worsening** |  |  |
| Indication: refractory headache<br>(vs. acute threat of visual deterioration) | Not calculable because no cases of visual worsening were observed in the refractory headache group. |  |  |
|  | **Headache improvement** |  |  |
| Indication: refractory headache<br>(vs. acute threat of visual deterioration) | 0.11 | 0.02 – 0.62 | 0.012 |
| Headache frequency at baseline<br>(per 5 MHD) | 0.75 | 0.63 – 0.89 | 0.002 |
|  | Pseudo-R-squared 0.387<br>Not retained in the model<sup>1</sup>: sex, age, time since diagnosis<br>BMI, CSF-OP |  |  |
|  | **Resolution of papilledema** |  |  |
| Indication: refractory headache<br>(vs. acute threat of visual deterioration) | Not calculable because no cases of no resolution of papilledema were observed in the refractory headache group. |  |  |
|  | **Loss of GCL volume** |  |  |
| Indication: refractory headache<br>(vs. acute threat of visual deterioration) | Not calculable because no cases of GCL volume loss were observed in the refractory headache group. |  |  |
|  | **Severe adverse event** |  |  |
| Indication: refractory headache<br>(vs. acute threat of visual deterioration) | 1.41 | 0.22 – 8.99 | 0.715 |
|  | Pseudo-R-squared 0.301<br>Not retained in the model<sup>2</sup>: sex, age, time since diagnosis |  |  |

calculated using binary-logistic bias-reduced logistic regression models according to Firth with clinical endpoints as the dependent variable and indication for surgical intervention as the independent variable (refractory headache vs. acute threat of visual deterioration as reference). Values above/below 1 indicate a higher/lower probability of clinical endpoints occurring with indication refractory headache compared to acute threat of visual deterioration.
<sup>1</sup>cAIC was used to select the best-fitting model for adjustment from a predefined set of known relevant covariables: sex, age, time since diagnosis, BMI, CSF-OP, MHD/headache severity at baseline and all other variables displaying an association with the respective outcome parameter $p < 0.2$ in univariable analyses.
<sup>2</sup>cAIC was used to select the best-fitting model for adjustment from a predefined set of known relevant covariables: sex, age, time since diagnosis and all other variables displaying an association with the respective outcome parameter $p < 0.2$ in univariable analyses.
BMI: Body Mass Index. cAIC: Corrected Akaike information criterion. CSF: cerebrospinal fluid. CSF-OP: CSF opening pressure. GCL: ganglion cell layer. MHD: monthly headache days. SAE: severe adverse event. VIF: variable inflation factor.

Multivariable analyses adjusting for relevant covariates showed that surgery indicated for refractory headache was associated with a significantly lower likelihood of achieving headache improvement (odds ratio 0.11, p=0.012) compared to surgery indicated for acute threat of visual deterioration, while the risk for SAEs did not differ between groups.

Again, goodness-of-fit of the multivariable models was moderate (Pseudo-R-squared 0.301-0.387), Hodges-Lehmann Gamma indicated robustness to bias by unidentified confounders.

For the other outcome measures, multivariable models could not be constructed because no cases of visual improvement, visual worsening, non-resolved papilledema, or GCL volume loss were observed in the refractory headache group (Table 3).

## Discussion

In this multi-center study derived from population-based data, we systematically evaluated the outcomes of surgical interventions in pwIIH and examined the influence of treatment indication on these outcomes. Our findings demonstrate that surgical intervention was only indicated in 3.6% of our cohort and that both CSF diversion procedures and ONSF are effective in cases of acute threat of visual deterioration, with significant improvements in visual function observed. However, resolution of papilledema occurred twice as often as visual improvement, suggesting that interventions were presumably performed when irreversible retinal damage had already occurred.

Still, further visual worsening after surgical intervention was noted in 13.9% – and only in association with adverse events resulting in insufficient ICP reduction – indicating that surgical interventions were effective if successful.

In contrast, when refractory headache was the sole indication for surgery, the likelihood of visual improvement was markedly reduced, however, vision was significantly milder affected in those. Multivariable analysis confirmed a significantly lower probability of headache improvement in this subgroup.

### Comparative Efficacy and Safety of CSF Diversion and ONSF

Our results corroborate previous reports that highlight the potential of surgical interventions to improve vision in IIH, although existing literature has been largely limited to case reports or single-center studies with small sample sizes and heterogeneous outcome measures [14, 15, 17, 18]. In contrast, our multi-center study provides a more robust analysis, offering a comprehensive multidisciplinary assessment both pre- and post-intervention, along with thorough adjustments for relevant confounders. While previous meta-analyses suggested differential efficacy and safety profiles with ONSF reported to have higher success rates regarding visual field changes, and CSF diversion to provide slightly better headache relief, our study found no significant differences in key outcomes, including visual improvement, visual worsening, papilledema resolution and headache improvement comparing CSF diversion and ONSF [14, 17, 26]. Thus, both approaches may be viable options when appropriately indicated. GCL volume loss, an objective marker of optic nerve damage, was comparable between CSF diversion (38.5%) and ONSF (33.3%) at 6 months follow-up. This is clinically particularly important as it provides an objective structural correlate to functional visual outcomes, reinforcing the comparable efficacy of both interventions.

Complication profiles of CSF diversion and ONSF were statistically indifferent regarding overall SAE (37.0 vs. 44.4%), as well as revision rates (34.6 vs. 44.4%). However, revision for failure to resolve papilledema was found exclusively in ONSF (33.3%), whereas revision for dysfunction occurred exclusively with CSF diversion (34.6%). This could be partially explained by ONSF only being performed by visual threat, whereas the CSF diversion procedures included patients of heterogeneous /varying degrees of visual compromise.

Previous literature has suggested that CSF diversion techniques are associated with a 9.4% severe complication rate and a 43.4% failure rate, compared to ONSF’s 2.2% severe complication rate and 9.4% failure rate [14, 15, 26]. This discrepancy may also reflect differences in definitions, follow-up periods, or patient selection criteria across studies and warrant proper randomized studies.

### The Critical Role of Treatment Indication

Arguably the most clinically relevant finding of our study is the substantial impact of treatment indication on outcomes. Our data demonstrate that, while interventions for acute threat of vision yielded favorable outcomes, surgical procedures performed solely for refractory headache not only failed to achieve visual improvement (0% vs. 50%) – as expected given the indication – but were also associated with a 10-fold lower likelihood of achieving headache improvement, which is concerning. This finding strongly suggests that contrary to earlier reports from two to three decades ago, refractory headache in the absence of visual deterioration may not be considered a valid standalone indication for surgical intervention in IIH [27, 28].

Thus, patient selection based on precise indications is crucial for optimizing outcomes in IIH management. The traditional view that surgical interventions can effectively address both visual deterioration and refractory headache in pwIIH requires reconsideration based on our results.

Of note, the overall proportion of pwIIH requiring surgical intervention in our cohort (3.6%) appears notably lower than previously reported in the literature, where contemporary reports from population-level data range from 9-20% [29, 30]. This may reflect the benefits of a comprehensive, interdisciplinary management approach offered at tertiary care centers. Early diagnosis, close monitoring, and individualized medical management by neurology, ophthalmology, and neurosurgery specialists likely contribute to more effective conservative treatment and a reduced need for surgical escalation.

### Alternative Management Strategies for Refractory Headache in IIH

Given our finding that surgical intervention appears inappropriate for pwIIH with refractory headache as the sole indication, it is important to consider alternative management strategies for this subgroup. Recent literature suggests that headache in IIH may persist in up to 50% of pwIIH even after resolution of papilledema and normalization of ICP, highlighting a complex and multifactorial pathophysiology that extends beyond elevated ICP alone [31].

Evidence for the alleviation of headache in IIH is much needed as it barely exists. Currently, consensus expert opinion is to treat the headache according to its phenotypic resemblance of primary headache disorders, but this is not evidence-based [5, 13]. Calcitonin gene-related peptide (CGRP) monoclonal antibodies, such as erenumab, may benefit pwIIH with persistent headaches despite resolved papilledema, although randomized trials are lacking [32–34]. In pwIIH with predominant headache symptoms, pharmacological and non-pharmacological approaches specifically targeting headache mechanisms within a comprehensive multidisciplinary setting may be more appropriate than surgical interventions aimed at reducing ICP [8–10].

### Strengths and Limitations

Our study has several strengths. Its multi-center design, grounded in population-based data, enhances the generalizability of our findings across diverse clinical settings. The use of comprehensive and standardized outcome measures, obtained within specialized multidisciplinary teams, ensures consistency in data collection and interpretation. Furthermore, the rigorous adjustment for relevant covariates and the explicit consideration of treatment indication as a potential predictor of outcomes add robustness to the analysis [35].

However, several limitations must be acknowledged. First, the retrospective nature of the study may have introduced selection bias, despite our efforts to include a well-defined cohort based on the revised Friedman criteria screening all pwIIH treated during the respective interval at all three centers. Specifically, there may be an inherent bias introduced by grouping pwIIH according to surgical indication, as those undergoing surgery for refractory headache were diagnosed with IIH for a longer duration prior to intervention and, consequently, likely had a higher baseline risk of chronification independent of the indication itself, but a lower degree of papilledema. Although we adjusted for this in the multivariable model, residual confounding cannot be entirely excluded. Second, despite the multi-center design, the relatively small sample size – particularly in the refractory headache subgroup – limited the statistical power of some analyses and precluded the construction of multivariable models for certain outcome measures. Third, the heterogeneity in surgical techniques and postoperative management across centers and time could have influenced the results. Additionally, the follow-up period of six months may not capture long-term outcomes and complications. Another important limitation is the lack of a control group or randomization, which precludes definitive causal inferences about the relative efficacy of different interventions. Treatment decisions were made in routine clinical practice and may have been influenced by unmeasured variables, introducing potential confounding. However, this is mitigated by the structure of the centers’ databases, which include most pwIIH from their respective geographical areas, and the very unselective inclusion criteria [35, 36]. Also, Rosenbaum sensitivity tests with Hodges-Lehmann Gamma indicated robustness to bias by unidentified confounders [24].

### Clinical Implications and Future Directions

Our findings have several important clinical implications for the management of pwIIH who are being considered for surgical intervention. First, our results strongly support a judicious, indication-based approach, reinforcing that surgical interventions should be reserved for the small subset of pwIIH with an acute threat of visual deterioration. In contrast, for pwIIH with predominantly refractory headache in the absence of visual compromise, alternative management strategies should be explored. These conclusions are aligned with current international consensus guidelines, which advocate that decisions regarding invasive IIH treatment indications should be determined by multidisciplinary specialized boards [6, 7, 10]. Importantly, our findings indicate that the choice between CSF diversion procedures and ONSF should be based less on assumptions of differential efficacy or safety, as both appear to offer comparable outcomes when appropriately indicated. Instead, factors such as anatomical feasibility, surgeon expertise, institutional protocols, and patient preferences should guide procedural selection.

Future research should focus on prospective, ideally randomized studies comparing different surgical approaches for IIH, with longer follow-up periods and larger sample sizes. However, such studies currently appear unfeasible due to the low number of pwIIH as well as the significant resources required and limited commercial interest in this field. The potential role of newer interventions, such as venous sinus stenting, particularly for pwIIH with refractory headache, deserves further investigation. Moreover, developing combination approaches or sequential treatment algorithms may be necessary for pwIIH presenting with both visual symptoms and refractory headache, recognizing that distinct mechanisms may underlie these distinct manifestations of IIH. In any case, although time since diagnosis was not significantly associated with outcome in our cohort, it seems likely that earlier surgical intervention, when applied to appropriately selected patients, may lead to improved clinical outcomes by minimizing neuroaxonal damage to the optic nerve and chronification of headache optimizing recovery potential.

In conclusion, this study confirms that CSF diversion procedures and ONSF are effective in improving visual outcomes – and to a much lesser extent headache – in pwIIH when performed due to an acute threat of visual deterioration. However, refractory headache alone does not constitute an appropriate indication for surgical intervention. These findings underscore the critical importance of careful patient selection and highlight the need for a tailored, indication-based approach to the surgical management of IIH. Further studies specifically targeting the optimal type and timing of surgical interventions are warranted.

## Funding

This study was partially funded by the Medical Scientific Fund of the Mayor of the City of Vienna.

## Acknowledgement

We thank all the VIIH investigators, clinical research staff, and especially the patients for helping to collect these data. The named individuals were not compensated for their help. <u>VIIH investigators in alphabetical order:</u> Bsteh, Gabriel (Department of Neurology, Medical University of Vienna); Kirchner, Karl (Department of Ophthalmology, Medical University of Vienna); Krajnc, Nik (Department of Neurology, Medical University of Vienna); Macher, Stefan (Department of Neurology, Medical University of Vienna); Michl, Martin (Department of Ophthalmology, Medical University of Vienna); Mitsch, Christoph (Department of Ophthalmology, Medical University of Vienna); Müller, Nina (Department of Neurology, Medical University of Vienna); Pemp, Berthold (Department of Ophthalmology, Medical University of Vienna); Reitner, Andreas (Department of Ophthalmology, Medical University of Vienna); Stapf, Christoph (Department of Ophthalmology, Medical University of Vienna); Wöber, Christian (Department of Neurology, Medical University of Vienna); Zebenholzer, Karin (Department of Neurology, Medical University of Vienna).

## Potential conflicts of interest

**Gabriel Bsteh:** has participated in meetings sponsored by, received speaker honoraria or travel funding from Biogen, Celgene/BMS, Janssen, Lilly, Medwhizz, Merck, Neuraxpharm, Novartis, Roche, Sanofi-Genzyme and Teva, and received honoraria for consulting Adivo Associates, Biogen, Celgene/BMS, Janssen, Merck, Novartis, Roche, Sanofi-Genzyme and Teva. He has received unrestricted research grants from Celgene/BMS and Novartis. He serves on the Executive Committee of the European Committee for Treatment and Research in Multiple Sclerosis (ECTRIMS) and the Board of Directors of the International Multiple Sclerosis VisualSystem Consortium (IMSVISUAL).

**Nadja Skadkær Hansen**: received Funding from Novo Nordisk Foundation and Migrænefonden Af 1988, a lecture fee from Pfizer, and a travel grant from Augustinusfonden during the conduction of this work.

**Sina Zaic**: declares no conflict of interest relevant to this study

**Steffen Hamann**: declares no conflict of interest relevant to this study.

**Johanne Juhl Korsbæk**: received funding from the Lundbeck Foundation (276-2018-40 and 380-2021-1140), Candys Foundation (2015-146), Odense University Hospital and Rigshospitalet (25-A1320, 69-A3346) during the conduct of the study.

**Nik Krajnc**: has participated in meetings sponsored by, received speaker honoraria or travel funding from Alexion, BMS/Celgene, Janssen-Cilag, Merck, Neuraxpharm, Novartis, Roche and Sanofi-Genzyme and held a grant for a Multiple Sclerosis Clinical Training Fellowship Programme from the European Committee for Treatment and Research in Multiple Sclerosis (ECTRIMS).

**Stefan Macher**: declares no conflict of interest relevant to this study.

**Laleh Dehghani Molander**: declares no conflict of interest relevant to this study.

**Klaus Novak**: declares no conflict of interest relevant to this study.

**Therese Wallentin Steenfoss**: declares no conflict of interest relevant to this study.

**Marianne Wegener**: declares no conflict of interest relevant to this study.

**Berthold Pemp**: has received honoraria for consultancy/speaking from Chiesi, GenSight and Santen

**Rigmor Hoejland Jensen**: has received grants from Københavns Universitet, Lundbeck Pharma, Novo Nordisk, and Lundbeck Foundation paid to the institution and serving as Chair of Master of Headache Disorders at University of Copenhagen and Director in Lifting The Global Burden of Headache (unpaid).

**Dagmar Beier:** has participated in meetings sponsored by, received speaker honoraria or travel funding from Abbvie, Allergan, Lundbeck, Novartis, Pfizer, and Teva, and received honoraria for consulting Abbvie, Lilly, Lundbeck, Pfizer and Teva. She has participated in clinical trials and/or received research grants for Lilly, Lundbeck, Novartis, Novo Nordic Foundation and Teva.

## Author contributions

**Gabriel Bsteh:** study concept and design, acquisition of data, statistical analysis and interpretation of data, drafting of manuscript

**Nadja Skadkær Hansen**: acquisition of data, interpretation of data, critical revision of manuscript for intellectual content

**Sina Zaic**: acquisition of data, critical revision of manuscript for intellectual content

**Steffen Hamann**: acquisition of data, critical revision of manuscript for intellectual content

**Johanne Juhl Korsbæk**: acquisition of data, critical revision of manuscript for intellectual content

**Nik Krajnc**: acquisition of data, critical revision of manuscript for intellectual content

**Stefan Macher**: acquisition of data, critical revision of manuscript for intellectual content

**Laleh Dehghani Molander**: acquisition of data, critical revision of manuscript for intellectual content

**Klaus Novak**: acquisition of data, critical revision of manuscript for intellectual content

**Therese Wallentin Steenfoss**: acquisition of data, critical revision of manuscript for intellectual content

**Marianne Wegener**: acquisition of data, critical revision of manuscript for intellectual content

**Berthold Pemp**: acquisition of data, critical revision of manuscript for intellectual content

**Rigmor Hoejland Jensen**: study concept and design, acquisition of data, interpretation of data, critical revision of manuscript for intellectual content, study supervision (contributed equally)

**Dagmar Beier:** study concept and design, acquisition of data, interpretation of data, critical revision of manuscript for intellectual content, study supervision (contributed equally)

## Notes

### Funding Statement

This study was partially funded by the Medical Scientific Fund of the Mayor of the City of Vienna (Project 21009).

### Author Declarations

The study was approved by the ethics committee of the Medical Universities of Vienna (ethics approval number: 2216/2020), the Region of Southern Denmark (journal number: 24/17408, approval covers all Danish sites) and the Danish Data Protection Agency (journal number: 24/21145). As this was a retrospective study, the need for written informed consent from study participants was waived by the ethics committee and the Region of Southern Denmark.

