## Supplemental Table 1 for "Surgical interventions in idiopathic intracranial hypertension – a comprehensive multi-center study of outcome and the role of treatment indication"

**Supplemental Table 1. Severe Adverse Events.**

**Abbreviations:** ONSF: optic nerve sheath fenestration. VA: ventriculo-atrial. VP: ventriculo-peritoneal.

| **ID** | **Intervention** | **Indication** | **SAE** |
| --- | --- | --- | --- |
| **C-01** | ONSF | Acute threat of vision | None |
| **C-02** | ONSF | Acute threat of vision | None |
| **C-03** | ONSF | Acute threat of vision | None |
| **C-04** | CSF diversion (VP shunt) | Acute threat of vision | Revision (VP shunt replaced by VA-shunt) |
| **C-05** | CSF diversion (VP shunt) | Acute threat of vision | Visual worsening, revision (due to blockage of drain system) |
| **C-06** | CSF diversion (VP shunt) | Acute threat of vision | Revision (peritoneal displacement) |
| **C-07** | ONSF | Acute threat of vision | None |
| **C-08** | ONSF | Acute threat of vision | Visual worsening, failure of ONSF (VP shunt inserted) |
| **C-09** | ONSF | Acute threat of vision | None |
| **C-10** | ONSF | Acute threat of vision | Drug induced liver injury (due to acetazolamide) |
| **O-01** | CSF diversion (VA shunt) | Acute threat of vision | None |
| **O-02** | CSF diversion (VP shunt) | Acute threat of vision | None |
| **O-03** | ONSF | Acute threat of vision | Visual worsening, acute failure of ONSF (VP shunt inserted) |
| **O-04** | CSF diversion (VP shunt) | Acute threat of vision | None |
| **O-05** | CSF diversion (VP shunt) | Acute threat of vision | Revision (due to shunt dysfunction) |
| **O-06** | CSF diversion (VP shunt) | Refractory headache | Headache relapse (no shunt dysfunction detected) |
| **O-07** | CSF diversion (VP shunt) | Acute threat of vision | None |
| **O-08** | CSF diversion (VP shunt) | Acute threat of vision | Revision (due to shunt dysfunction) |
| **O-09** | CSF diversion (VP shunt) | Acute threat of vision | None |
| **O-010** | CSF diversion (VP shunt) | Acute threat of vision | None |
| **O-011** | CSF diversion (VP shunt) | Acute threat of vision | Revision (2x, ultimately replaced by VA-shunt) |
| **O-012** | ONSF | Acute threat of vision | Visual worsening, failure of ONSF after 1 year (VP shunt inserted) |
| **V-01** | CSF diversion (VP shunt) | Refractory headache | Revision (due to shunt dysfunction) |
| **V-02** | CSF diversion (VP shunt) | Acute threat of vision | Revision (due to shunt dysfunction) |
| **V-03** | CSF diversion (VP shunt) | Refractory headache | None |
| **V-04** | CSF diversion (VP shunt) | Refractory headache | None |
| **V-05** | CSF diversion (VP shunt) | Acute threat of vision | None |
| **V-06** | CSF diversion (VP shunt) | Acute threat of vision | None |
| **V-07** | CSF diversion (VP shunt) | Acute threat of vision | None |
| **V-08** | CSF diversion (VP shunt) | Acute threat of vision | None |
| **V-09** | CSF diversion (VP shunt) | Acute threat of vision | None |
| **V-10** | CSF diversion (VP shunt) | Acute threat of vision | Visual worsening, revision due to shunt infection (3x, ultimately replaced by VA-shunt) |
| **V-11** | CSF diversion (VA shunt) | Refractory headache | None |
| **V-12** | CSF diversion (VP shunt) | Acute threat of vision | None |
| **V-13** | CSF diversion (VP shunt) | Refractory headache | None |
| **V-14** | CSF diversion (VP shunt) | Acute threat of vision | None |
