## Supplemental Table 2 for "Surgical interventions in idiopathic intracranial hypertension – a comprehensive multi-center study of outcome and the role of treatment indication"

**Supplemental Table 2. Baseline characteristics stratified by type of surgical intervention**

**Abbreviations:** BMI: Body mass index. CSF-OP: cerebrospinal fluid opening pressure. dB: decibels. GCL: ganglion cell layer. logMAR: logarithm of the minimum angle of resolution. ONSF: optic nerve sheath fenestration. pRNFL: peripapillary retinal nerve fiber layer. NRS: numerical rating scale.

^1^absolute number and percentage. ^2^mean and standard deviation. ^3^median and inter-quartile range. ^4^independent t-test. ^5^Mann-Whitney U test.

|  | **CSF diversion**  (n=27) | **ONSF**  (n=9) | **p-value** |
| --- | --- | --- | --- |
| Females^1^ | 27 (100) | 9 (100) | n.a. |
| Age at diagnosis^2^ (years) | 32.1 (8.0) | 33.6 (9.4) | 0.647^4^ |
| Time since diagnosis^3^ (months) | 1.7 (0.1 – 23.6) | 0.3 (0.1 – 1.0) | 0.180^5^ |
| BMI^3^ | 35.7 (33.1 – 44.5) | 37.5 (28.2 – 46.9) | 0.818^5^ |
| CSF-OP^3^ (cm H2O) | 38 (29.5 – 49.5) | 45.0 (40.5 – 50.0) | 0.195^5^ |
| Visual impairment^1^ | 23 (85.2) | 8 (88.9) | 0.999^4^ |
| Best corrected visual acuity^3^ (logMAR, worse eye) | 0.10 (0 – 0.40) | 0.26 (0.13 – 0.37) | 0.293^5^ |
| Perimetric mean defect^2^ (dB, worse eye) | -7.1 (-22.8 – -2.7) | -16.1 (-25.8 – -6.2) | 0.579^5^ |
| Frisén grade^3^ | 3 (2 – 4) | 3 (1 – 4) | 0.214^5^ |
| pRNFL thickness^2^ (µm, worse eye) | 195 (125) | 231 (237) | 0.673^4^ |
| GCL volume^2^ (mm^3^, worse eye)* | 0.93 (0.74) | 1.09 (0.95) | 0.129^4^ |
| Headache frequency^3^ (days per month) | 30 (30 – 30) | 30 (19.5 – 30) | 0.720^5^ |
| Headache severity^3^ (NRS) | 8 (7 – 10) | 8 (8 – 10) | 0.856^5^ |
| Acetazolamide dosage^3^ (mg/day) | 1500 (1000 – 2000) | 1750 (1000 – 3000) | 0.485^5^ |
| Number of lumbar punctures before surgery^3^ | 2 (1 – 5) | 1 (1 – 4) | 0.377^5^ |
